# NeoCVST Score: Classification of Brain Injury Secondary to Neonatal Cerebral Venous Sinus Thrombosis

**DOI:** 10.64898/2026.05.06.26352611

**Authors:** Rhandi Christensen, Linda S. de Vries, Mehmet N. Cizmeci, Pradeep Krishnan, Vann Chau, Nomazulu Dlamini, Elizabeth Pulcine, Mahendra Moharir

## Abstract

**Background:** Neonatal cerebral venous sinus thrombosis (CVST) is associated with intracranial hemorrhage (ICH) and ischemic lesions. There is no scale to characterize the spectrum of brain injury secondary to neonatal CVST.

**Objective:** To develop the Neonatal CVST Hemorrhage Score (NeoCVST Score) to characterize ICH and brain injury in neonates with CVST.

**Methods:** This was a retrospective cohort study of neonates with CVST diagnosed using brain MRI/MRV. The NeoCVST Score was developed using the study cohort, integrating elements from previous hemorrhage classification systems and expert consensus. Logistic regression examined associations between NeoCVST score and neurodevelopmental outcomes (Pediatric Stroke Outcome Measure). Interrater reliability was assessed with intraclass correlation coefficient.

**Results:** The study included 100 neonates (77% term and 23% preterm) with CVST. Thrombosis of multiple venous sinuses was present in 62%. ICH was present in 63%. Supratentorial hemorrhage was present in 57% and included germinal matrix hemorrhage and intraventricular hemorrhage (GMH-IVH) grades 1-2 (22%), GMH-IVH grade 3 (15%), parenchymal (43%) and thalamic (18%) hemorrhage. Infratentorial hemorrhage was present in 19% and included cerebellar (18%) and brainstem (4%) hemorrhage. Extra-axial hemorrhage was present in 32% and included epidural (2%), subdural (26%) and subarachnoid hemorrhage (6%). Ischemic brain injury was present in 67% and included lesions in the medullary vein distribution (13%), white matter (54%), basal ganglia (17%) and thalamus (25%). Neurodevelopmental outcomes included 40% with normal outcomes and 60% with neurodevelopmental impairments. NeoCVST total score (OR=1.1, P=0.02) and subscores for thalamic hemorrhage (OR=1.9, P=0.04), thalamic ischemia (OR=2.2, P=0.005) and bilateral thalamic ischemia (OR=2.8, P=0.01) were predictors of adverse neurodevelopmental outcome. Inter-rater reliability showed moderate-good agreement between reviewers with an intraclass correlation coefficient of 0.71.

**Conclusions:** The NeoCVST Score is a simple clinical tool to characterize ICH and brain injury secondary to neonatal CVST. Increasing NeoCVST total score and subscores for thalamic hemorrhage and ischemia were associated with worse neurodevelopmental outcomes.

## Introduction

Cerebral venous sinus thrombosis (CVST) is a thrombotic occlusion of cerebral veins and dural sinuses. The incidence of neonatal CVST has been reported to be as high as 1 per 9100 neonates, which is higher than the incidence of CVST in children and adults^1,2^. Risk factors for CVST include perinatal factors (vacuum or forceps assisted delivery), and neonatal factors (dehydration, infection, surgery)^1,3,4^. Neonates with CVST may be asymptomatic or present with seizures and encephalopathy^1,4^. CVST can be diagnosed with brain magnetic resonance imaging (MRI) and magnetic resonance venography (MRV) or computed tomography (CT) and CT cerebral venography (CTV). Neonatal CVST can cause venous congestion and lead to parenchymal brain injury and multi-compartmental hemorrhage. This extensive brain injury secondary to neonatal CVST is associated with an increased risk of mortality and long-term neurological impairments^3–5^. Intracranial hemorrhage (ICH) and ischemic brain injury secondary to neonatal CVST can be highly variable and can differ in location, size and severity^3,5,6^.

There is currently no classification system to characterize ICH and brain injury secondary to neonatal CVST and to predict outcomes. ICH classification systems that have been developed for adult and pediatric populations do not adequately describe hemorrhage due to neonatal CVST^7–9^. The Pediatric Intracerebral Hemorrhage (Pediatric ICH) Score and the Modified Pediatric ICH score do not include the location and type of hemorrhage, and estimating the size of hemorrhage as total brain volume percentage can be challenging^7,10^. The Periventricular Hemorrhage Infarction (PVHI) Score is limited to hemorrhage from PVHI in infants born preterm^11^. Hong and Lee described neonatal ICH based on location and type of hemorrhage, however there is no numerical score to grade the severity of hemorrhage^8^. The Pediatric Intracerebral Hemorrhage (PIH) Score grades the severity of hemorrhage in pediatric populations, but does not describe the hemorrhage. The Neonatal Intracerebral Hemorrhage (NIH) Classification was developed to characterize hemorrhage in neonates based on location^8^. However, the NIH Classification does not grade the severity or include high risk features. Prior ICH classifications systems do not consider concurrent ischemic brain injury which can also influence management and prognostication.

A classification system for neonatal CVST related ICH is needed to describe and grade the severity of hemorrhage, guide neuroprognostication, and help with treatment decision-making. In clinical practice, the role of anticoagulation in treating neonatal CVST is unclear, and some centres proceed with anticoagulation in the presence of ICH, while other centres are hesitant to treat in the presence of ICH. Previous observational cohort studies have demonstrated the safety of anticoagulation treatment in neonatal CVST^5,12,13^. Despite these encouraging findings, the use of anticoagulation for neonatal CVST remains mixed. A neonatal CVST brain injury score could discriminate between mild and severe brain injury and define inclusion/exclusion criteria for randomized controlled trials examining the efficacy of anticoagulation in neonatal CVST. The primary objective of this study was to develop a standardized classification and scoring system for brain injury secondary to neonatal CVST (Neonatal CVST Hemorrhage Score or NeoCVST Score). Our secondary objective was to examine the associations between NeoCVST total score and subscores with neurodevelopmental outcomes. We hypothesized that increasing NeoCVST total score would be associated with worse neurodevelopmental outcomes.

## Methods

### Study Participants

We performed a retrospective chart review of neonates with radiologically confirmed CVST (MRI/MRV) admitted to The Hospital for Sick Children (Toronto, Canada) between January 1, 2000, to August 1, 2025. Study participants were identified from the Institutional Stroke Registry Database in the Children’s Stroke Program. Informed consent to participate in the Stroke Registry Database was obtained from the parent or legal guardian. The study was approved by the institutional research ethics board (REB #2663). A total of 148 neonates in the SickKids Stroke Registry with neonatal CVST were screened. Participants were included in the study if they were under 28 days old at the time of CVST diagnosis and had radiologically confirmed CVST (on MRI/MRV). Participants were excluded if they had vascular malformations (e.g. Vein of Galen malformation), or if thrombus was not confirmed on MRI/MRV (e.g. scans included cranial ultrasound or CT/CTV). Electronic charts were reviewed to collect clinical characteristics of study participants (sex, gestational age, perinatal history, comorbidities, age at CVST diagnosis, anticoagulation treatment, treatment complications, prothrombotic workup). Neurodevelopmental outcome was determined from Pediatric Stroke Outcome Measure (PSOM) score at the last Neurology clinic visit, and dichotomized as no functional impairment or normal outcome (PSOM total score < 0.5), or neurodevelopmental impairment (PSOM total score >0.5)^14^.

### Neuroimaging Data

Neuroimaging data was collected from MRI/MRV scans and radiology reports of neonates with CVST. The first MRI scan to detect CVST was reviewed by a Pediatric Neurologists (RC, MM) and Pediatric Neuroradiologist (PK) to determine thrombus characteristics, presence and severity of ICH and other parenchymal brain injury. Brain MRI axial sequences included anatomical (T1 and T2-weighted), diffusion weighted imaging (DWI), susceptibility weighted imaging (SWI) or multi-planar gradient recall (MPGR), and magnetic resonance venography (MRV). These sequences were used to determine the presence of CVST, ischemic brain injury (parenchymal lesions on DWI), intracranial hemorrhage (hemorrhage seen as T1 hyperintense signal or T2-hypointense signal, SWI or MPGR blooming) and any other brain lesion or malformation.

### Neonatal CVST Hemorrhage Score (NeoCVST Score)

We determined the predominant patterns of ICH and brain injury in our sample of neonates with CVST. We used this clinical data, along with previously published pediatric hemorrhage scoring systems^7,8,10,11^, to develop a classification system (NeoCVST Score) for brain injury secondary to neonatal CVST (Table 1). The classification system was developed with expert consensus from team members in Neurology (RC, VC, LD, MM), Neonatology (MC), and Neuroradiology (PK). All neonates in the study cohort were assessed using the NeoCVST Score by Pediatric Neurologists (RC, MM) and Pediatric Neuroradiologist (PK). ICH was characterized based on the specific features and assigned a numerical score for increasing severity as described below.

**Table 1.**
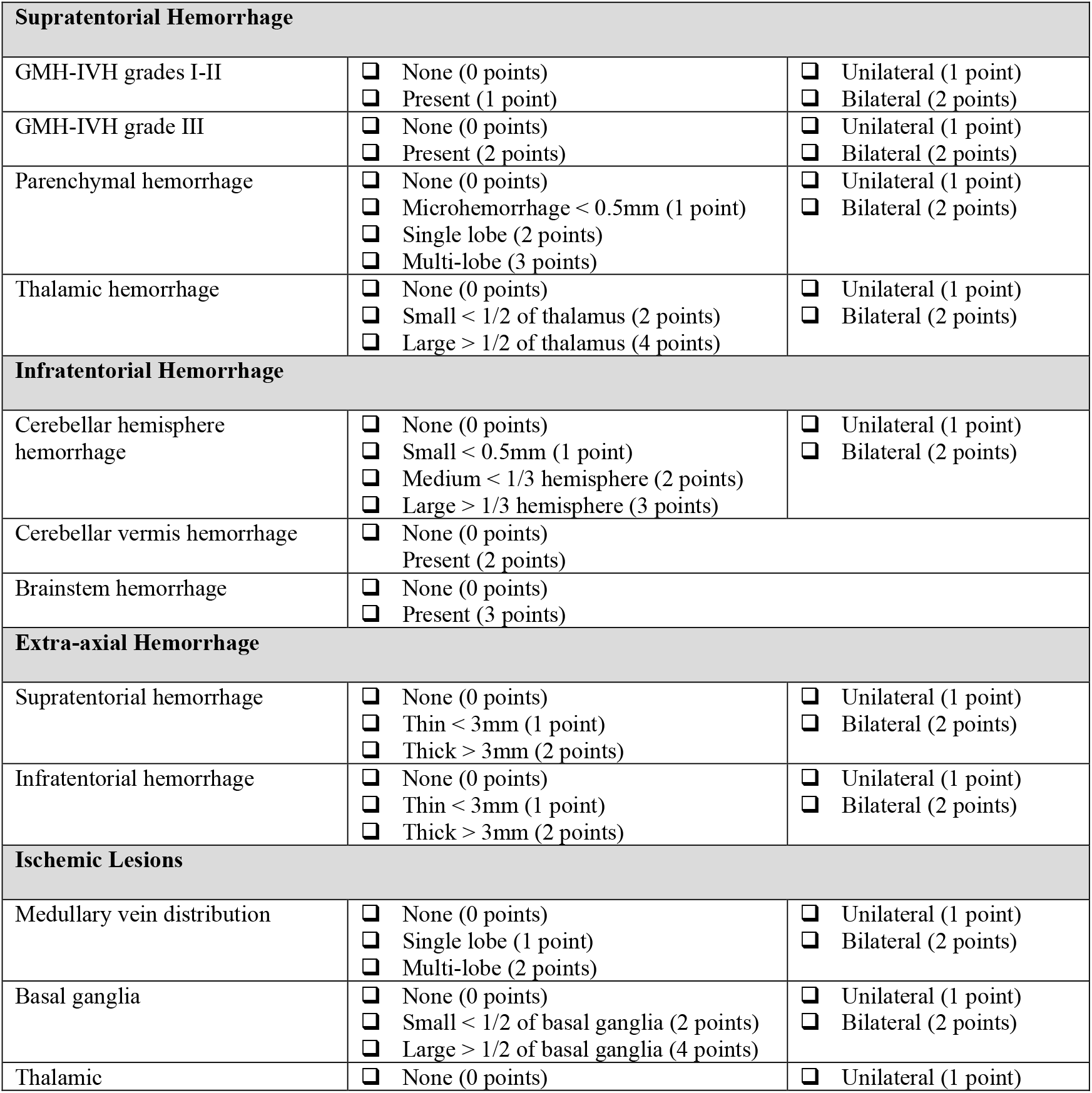
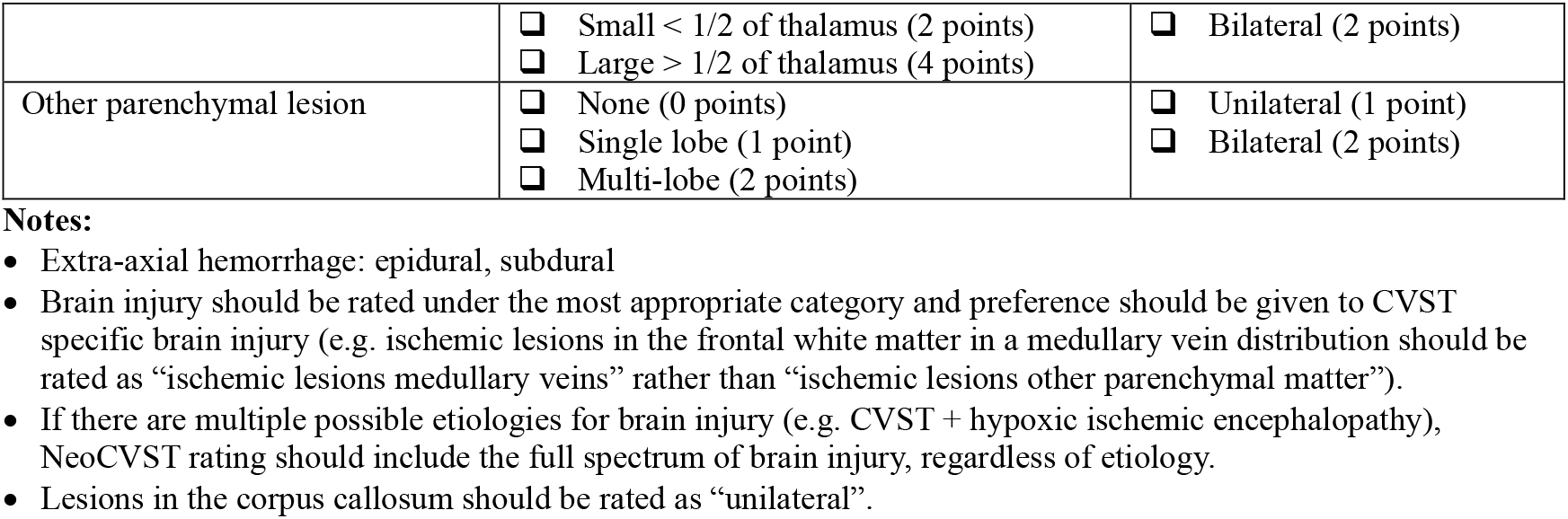
Neonatal CVST Score (NeoCVST Score)

- Location of hemorrhage

° Supratentorial
° Infratentorial
° Extra-axial (epidural, subdural, subarachnoid)

- Type of hemorrhage

° Germinal matrix hemorrhage and intraventricular hemorrhage (GMH-IVH)
° Parenchymal hemorrhage
° Specific structures: thalamus, cerebellum, brainstem

- Size

° Microhemorrhage
° Single lobe or multi-lobe
° Small or large relative to structure (e.g. thalamus, cerebellum)
° Thin or thick (extra-axial hemorrhage)

- Ischemic brain injury

° Medullary vein distribution
° White matter lesions
° Basal ganglia
° Thalamic

- Side

° Unilateral
° Bilateral

## Statistical Analysis

Statistical analyses were conducted using Stata version 18. Descriptive statistics were used to examine clinical characteristics, radiological features, treatment with anticoagulation, and neurodevelopmental outcome. Univariable and multivariable logistic regression were used to examine the association between NeoCVST total score and subscores with PSOM neurodevelopmental outcome (impairment vs no impairment as defined above). Interrater reliability for NeoCVST total score was assessed using a two-way mixed effects intraclass correlation coefficient for ratings by RC and PK.

## Results

A total of 148 neonates with CVST were screened from the Stroke Registry. From this, 48 neonates were excluded for not having MRI/MRV for CVST diagnosis, having a presumed thrombus which was not captured on imaging, and for having venous malformations. The final study sample included 100 neonates with CVST, and clinical characteristics are presented in Table 2. The study cohort included a greater proportion of males (79%). Most neonates were born at term (77%), and a smaller number were born preterm (23%). Perinatal risk factors for CVST included neonatal surgery (18%), vacuum or forceps assisted delivery (18%), hypoxic-ischemic encephalopathy (16%), congenital heart disease (11%), infection (10%), and coagulopathy (7%). Most neonates were symptomatic at the time of diagnosis with seizures (62%), and/or an abnormal neurological examination (38%).

**Table 2.** Clinical characteristics of neonates with CVST (N=100)

| Variable | Frequency (%) or Mean (SD) |
| --- | --- |
| Sex | Males: 79 (79%)<br>Females: 21 (21%) |
| Preterm and Term | Preterm: 23 (23%)<br>Term: 77 (77%) |
| Mean gestational age in weeks (SD) | 37.2 (3.3) |
| Perinatal risk factors | Vacuum or forceps assisted delivery: 18 (18%)<br>Surgery: 18 (18%)<br>Hypoxic ischemic encephalopathy: 16 (16%)<br>Congenital heart disease: 11 (11%)<br>Infection: 10 (10%)<br>Dehydration: 10 (10%)<br>Coagulopathy: 9 (9%) |
| Mean age at CVST diagnosis in days (SD) | 8.7 (6.7) |
| Clinical features at diagnosis | Seizures: 62 (62%)<br>Abnormal neurological exam: 38 (38%)<br>Asymptomatic: 18 (18%) |
| Location of thrombus | Multiple sinuses involved: 62 (62%)<br>Transverse sinus: 19 (19%)<br>Superior sagittal sinus: 15 (15%) |
| Intracranial hemorrhage<sup>b</sup> | Any hemorrhage 63 (63%)<br>Supratentorial hemorrhage: 57 (57%)<br>○ GMH-IVH grade 1-2: 22 (22%)<br>○ GMH-IVH grade 3: 15 (15%)<br>○ Parenchymal: 43 (43%)<br>○ Thalamus: 18 (18%)<br>Infratentorial hemorrhage: 19 (19%)<br>○ Cerebellum: 18 (18%)<br>○ Brainstem: 4 (4%)<br>Extra-axial hemorrhage 32 (32%)<br>○ Epidural: 2 (2%)<br>○ Subdural: 26 (26%)<br>○ Subarachnoid: 6 (6%) |
| Ischemic brain injury<sup>c</sup> | Ischemic lesions: 67 (67%)<br>Medullary vein distribution: 13 (13%) |
|  | White matter: 54 (54%)<br>Cystic white matter lesion: 4 (4%)<br>Basal ganglia: 17 (17%)<br>Thalamus: 25 (25%) |
| Mean NeoCVST Score | 9.4 (7.6) |
| Anticoagulation received | No: 34 (34%)<br>Yes: 58 (58%) at diagnosis, 8 (8%) after thrombus propagation |
| Mean duration of anticoagulation in months (SD) | 2.8 (1.2) |
| Reason for not anticoagulating (N=34) | Intracranial hemorrhage: 27 (80%)<br>○ GMH-IVH grade 1-2: 8 (24%)<br>○ GMH-IVH grade 3: 8 (24%)<br>○ Medullary vein: 6 (18%)<br>○ Parenchymal: 14 (41%)<br>○ Thalamic: 4 (12%)<br>○ Brainstem: 1 (3%)<br>○ Cerebellum: 5 (15%)<br>○ Extra-axial: 10 (29%)<br>Non-occlusive thrombus: 3 (8%)<br>Prematurity: 2 (6%)<br>Other: 2 (6%) |
| Worsening intracranial hemorrhage with anticoagulation (N=66) | 1 (2%) |
| Recanalization outcome (N=95) | Full recanalization: 48 (50%)<br>Partial recanalization: 41 (43%)<br>No change: 6 (6%) |
| Mean age at follow-up in years (SD) | 5.8 (4.9) |
| Mean PSOM Score at last follow-up (SD) | 1.3 (1.9) |
| Clinical outcome based on PSOM (N=94) | No impairment: 38 (40%)<br>Impairment: 56 (60%) |

<sup>a</sup>Cause of death: respiratory failure 3 (3%), sepsis 2 (2%), multiorgan failure 1 (1%), leukemia 1 (1%)
<sup>b</sup>Intracranial hemorrhage: some neonates had more than one type of intracranial hemorrhage injury
<sup>c</sup>Parenchymal brain injury: some neonates had more than one type of parenchymal injury

Neuroimaging findings for the cohort are presented in Table 2. The initial diagnostic scan was used to classify ICH in neonates with CVST. Most neonates had CVST involving multiple venous sinuses (62%). Supratentorial hemorrhage was present in 57% and included GMH-IVH grades 1-2 (22%), GMH-IVH grade 3 (15%), and parenchymal (43%) and thalamic (18%) hemorrhage. Infratentorial hemorrhage was present in 19% and included cerebellar (18%) and brainstem hemorrhage (4%). Extra-axial hemorrhage was present in 32% and included epidural (2%), subdural (26%) and subarachnoid hemorrhage (6%). Ischemic brain injury was present in 67% and included lesions in the medullary vein distribution (13%), white matter (54%), basal ganglia (17%) and thalamus (25%). Examples of ICH are presented in Figure 1, and examples of associated brain injury are presented in Figure 2. Follow-up imaging for 95 participants was available and showed full recanalization in 50%, partial recanalization in 44% and no change in 6%. A large proportion of neonates were treated with anticoagulation (66%) for a mean duration of 2.8 months. Of those that were not anticoagulated (N=34), reasons for not anticoagulating included the presence of ICH (80%), non-occlusive thrombus (8%), prematurity (6%) and other/not-specified (6%). Of the participants that received anticoagulation, most (98%) had no anticoagulation related complications such as new or worsening hemorrhage. One participant had worsening of extra-axial hemorrhage soon after starting anticoagulation therapy, therapy was stopped, and then restarted three days later with no further complications.

**Figure 1.**
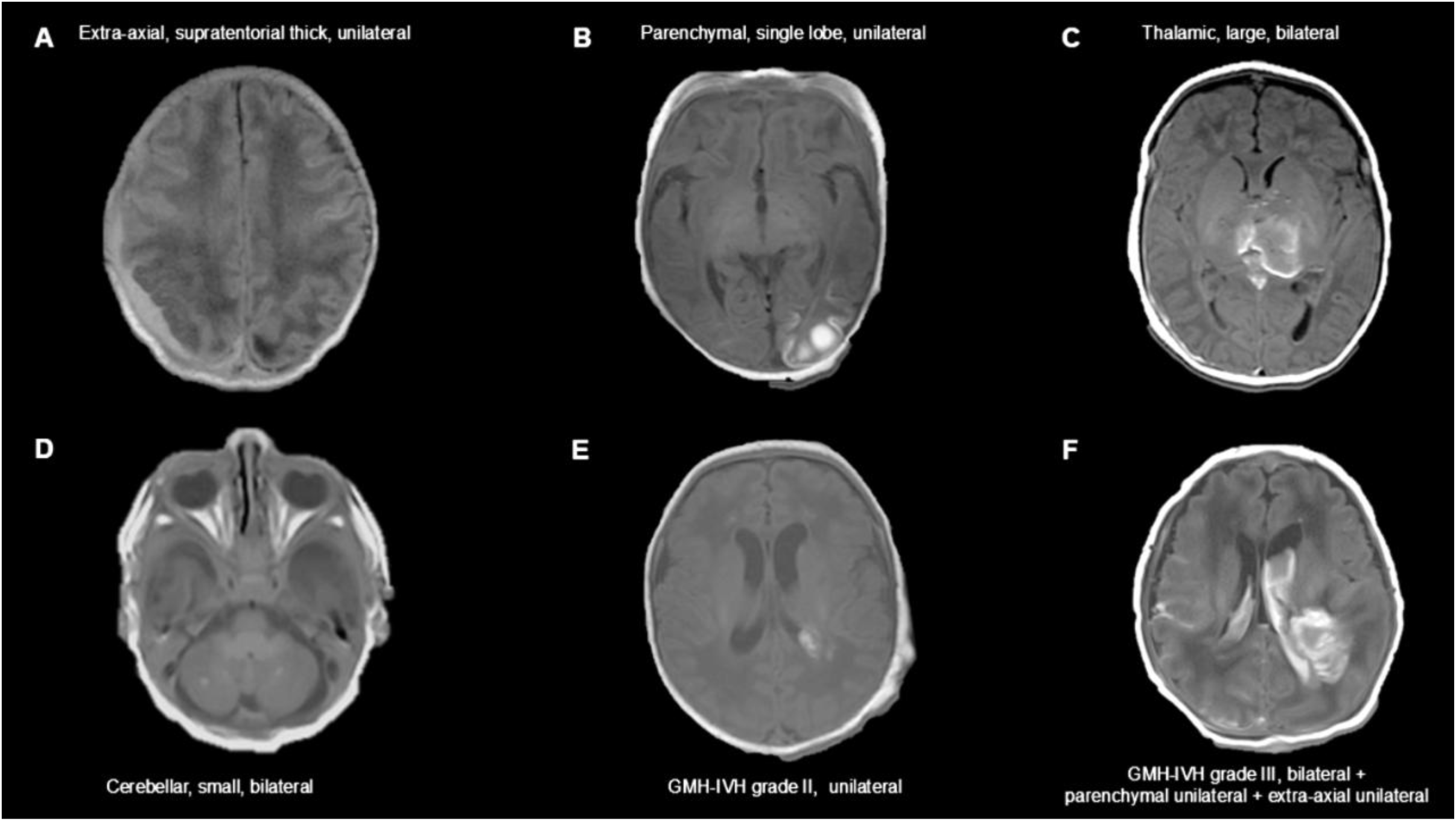
Spectrum of Intracranial Hemorrhage Secondary to Neonatal CVST.

**Figure 2.**
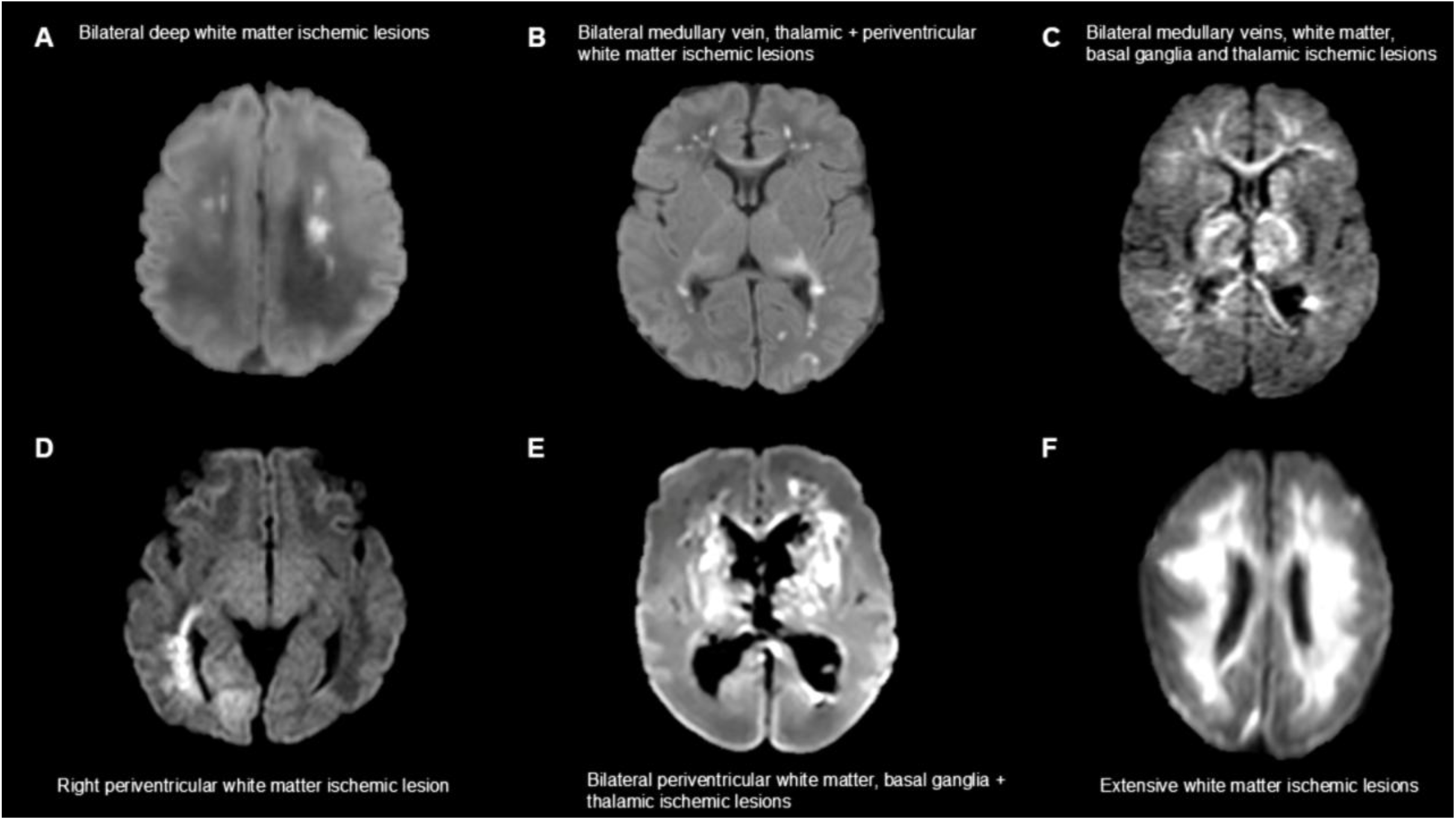
Spectrum of Brain Injury Associated with Neonatal CVST.

Clinical outcomes included a mortality rate of 5%, although none of the 5 deaths were related to CVST. Neurodevelopmental outcome based on PSOM score at the last clinical encounter was available for 95 participants (mean age at follow-up 5.8 years, SD 4.9). Normal outcomes were present in 40% and neurodevelopmental impairment was present in 60%.

The maximum NeoCVST total score is 85. The mean NeoCVST total score for the study cohort was 9.4 (SD: 7.6). Example scoring using the NeoCVST is presented in Figure 3. Intraclass correlation coefficient was 0.71 (95% CI: 0.12-1.00) indicating good agreement between the two raters. Logistic regression models were used to examine the association between NeoCVST total score and subscores with PSOM neurodevelopmental outcome (Table 3). Results from univariate regression models revealed that NeoCVST total score (OR=1.1, P=0.03) and subscores for thalamic hemorrhage (OR=1.9, P=0.03), thalamic ischemia (OR=2.1, P=0.009) and bilateral thalamic ischemia (OR=2.6, P=0.02) were predictors of neurodevelopmental outcome. These remained significant predictors after controlling for sex, prematurity, multi-focal hemorrhage and hypoxic-ischemic encephalopathy: NeoCVST total score (OR=1.1, P=0.02) and subscores for thalamic hemorrhage (OR=1.9, P=0.04), thalamic ischemia (OR=2.2, P=0.005) and bilateral thalamic ischemia (OR=2.8, P=0.01).

**Table 3.** Logistic Regression Models of NeoCVST Score and PSOM Clinical Outcomes.

| Univariate Analysis | Odds ratio | 95% CI | P-Value |
| --- | --- | --- | --- |
| NeoCVST total score | 1.1 | [0.9, 1.1] | 0.03* |
| NeoCVST subscore |  |  |  |
| – Supratentorial hemorrhage GMH-IVH grade 1-2 | 0.7 | [0.3, 2.0] | 0.5 |
| – Supratentorial hemorrhage GMH-IVH grade 1-2, bilateral | 0.9 | [0.5, 1.5] | 0.6 |
| – Supratentorial hemorrhage GMH-IVH grade 3 | 4.1 | [0.9, 4.6] | 0.07 |
| – Supratentorial hemorrhage GMH-IVH grade 3, bilateral | 2.1 | [1.3, 4.0] | 0.07 |
| – Supratentorial parenchymal hemorrhage | 1.3 | [0.9, 1.9] | 0.1 |
| – Supratentorial parenchymal hemorrhage, bilateral | 1.3 | [0.8, 2.1] | 0.4 |
| – Supratentorial thalamic hemorrhage | 1.9 | [1.1, 2.1] | 0.03* |
| – Supratentorial thalamic hemorrhage, bilateral | 2.6 | [0.9, 7.1] | 0.07 |
| – Infratentorial cerebellar hemisphere hemorrhage | 0.8 | [0.3, 2.0] | 0.7 |
| – Infratentorial cerebellar hemisphere hemorrhage, bilateral | 0.9 | [0.4, 1.8] | 0.8 |
| – Infratentorial cerebellar vermis hemorrhage | 1.1 | [0.3, 4.0] | 0.8 |
| – Infratentorial brainstem hemorrhage | 1.3 | [0.6, 2.8] | 0.5 |
| – Extra-axial hemorrhage, supratentorial | 0.6 | [0.3, 1.3] | 0.2 |
| – Extra-axial hemorrhage, supratentorial, bilateral | 0.8 | [0.4, 1.5] | 0.5 |
| – Extra-axial hemorrhage, infratentorial | 0.7 | [0.3, 1.5] | 0.4 |
| – Extra-axial hemorrhage, infratentorial, bilateral | 0.8 | [0.4, 1.4] | 0.1 |
| – Ischemic medullary vein distribution | 1.2 | [0.7, 2.4] | 0.5 |
| – Ischemic medullary vein distribution, bilateral | 1.2 | [0.6, 2.3] | 0.6 |
| – Ischemic white matter | 1.3 | [0.9, 2.1] | 0.2 |
| – Ischemic white matter, bilateral | 1.5 | [0.9, 2.3] | 0.09 |
| – Ischemic basal ganglia | 1.4 | [0.8, 2.3] | 0.2 |
| – Ischemic basal ganglia, bilateral | 1.6 | [0.8, 3.2] | 0.2 |
| – Ischemic thalamus | 2.1 | [1.2, 3.5] | 0.009* |
| – Ischemic thalamus, bilateral | 2.6 | [1.2, 5.6] | 0.02* |
| – Extra-axial hemorrhage, infratentorial, bilateral | 0.8 | [0.4, 1.4] | 0.1 |
| **Multivariate Analysis** (Covariates: preterm, sex, multi-focal hemorrhage, hypoxic-ischemic encephalopathy) | **Odds ratio** | **95% CI** | **P-Value** |
| NeoCVST total score | 1.1 | [1.0, 1.2] | 0.02* |
| NeoCVST Subscore |  |  |  |
| – Supratentorial thalamic | 1.9 | [1.0, 3.4] | 0.04* |
| – Ischemic thalamic | 2.2 | [1.3, 3.8] | 0.005* |
| – Ischemic thalamic, bilateral | 2.8 | [1.3, 6.3] | 0.01* |

**Figure 3.**
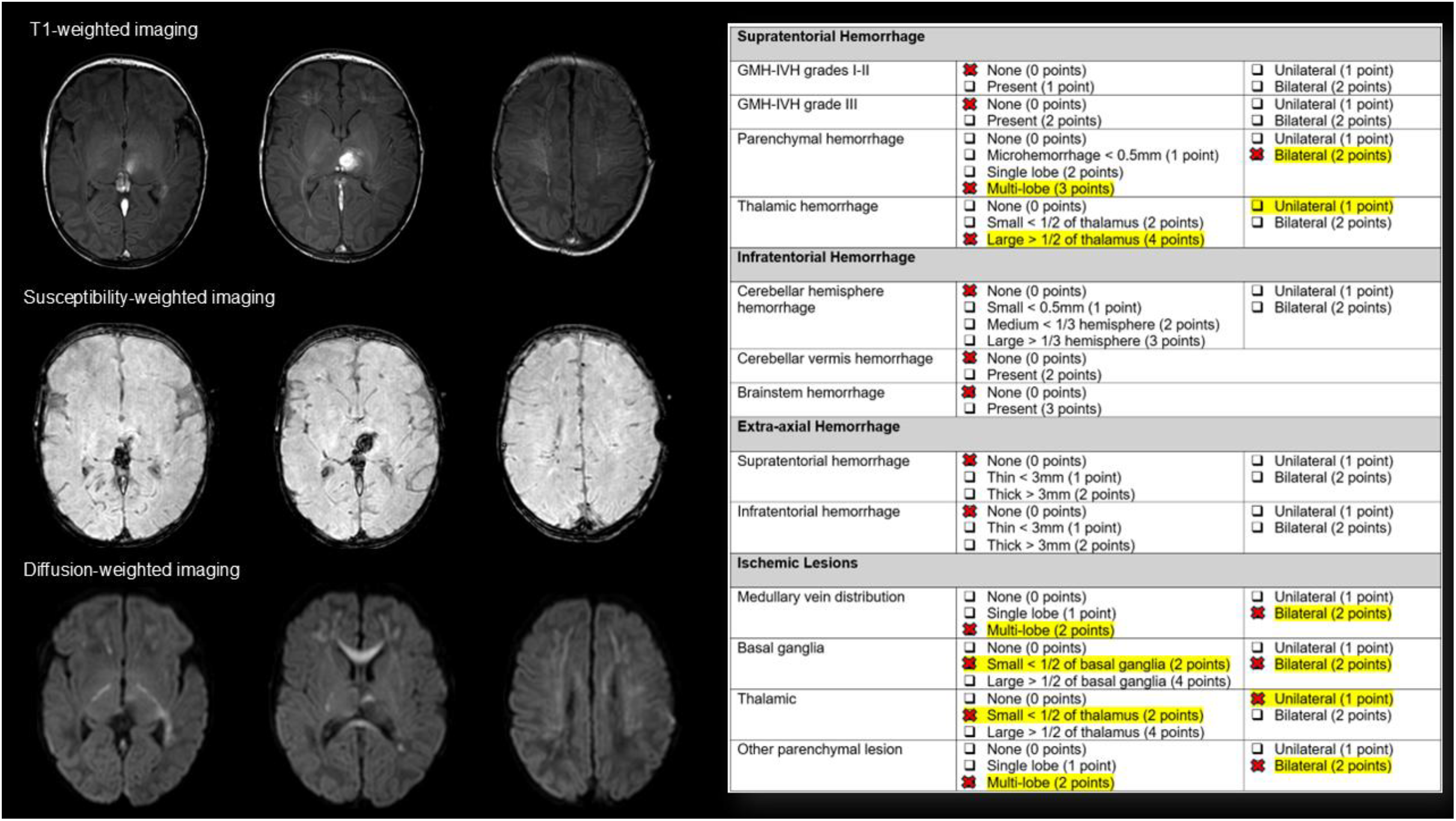
Example Scoring with NeoCVST Score.

Regression models were repeated with term neonates only (excluding preterm neonates, see supplemental table) and results were similar. Results from univariate regression models revealed that NeoCVST total score (OR=1.1, P=0.03) and subscores for thalamic hemorrhage (OR=2.1, P=0.04), bilateral thalamic hemorrhage (OR=4.7, P=0.04), thalamic ischemia (OR=3.2, P=0.003) and bilateral thalamic ischemia (OR=5.2, P=0.005) were predictors of neurodevelopmental outcome. These remained significant predictors after controlling for sex, multi-focal hemorrhage and hypoxic-ischemic encephalopathy: NeoCVST total score (OR=1.1, P=0.03) and subscores for thalamic hemorrhage (OR=2.1, P=0.04), bilateral thalamic hemorrhage (OR=4.7, P=0.04), thalamic ischemia (OR=3.2, P=0.003) and bilateral thalamic ischemia (OR=5.2, P=0.005).

## Discussion

We present the NeoCVST Score, a classification and scoring system for ICH and brain injury secondary to neonatal CVST. Neonatal CVST is associated with a spectrum of hemorrhagic brain injury and ischemic brain lesions which can complicate initial management decisions, and lead to long term morbidity and disability. We addressed an important clinical need with the development of the NeoCVST Score which can standardize the description and grading of the severity of ICH and brain injury in neonates with CVST.

Existing pediatric ICH classification systems could not adequately characterize hemorrhage in neonatal CVST and do not consider concurrent ischemic lesions. The Pediatric ICH Score by Beslow et al., is a simple grading scale of ICH for neonates and children aged 0-18 years old^7^. The positive features of the Pediatric ICH are the inclusion of high risk features, and a numerical score reflecting severity, with higher scores being associated with worse outcomes. The main limitation of this tool is that total brain volume is difficult for clinicians to measure and can be subjective. Furthermore, the Pediatric ICH does not characterize the type or location of hemorrhage, which is important as thalamic hemorrhage may have different clinical implication than a similar sized subdural hemorrhage. The Pediatric ICH score was modified by Guedon et al., to account for additional high risk features such as infratentorial hemorrhage^10^. However, this scale does not account for the type of hemorrhage, and estimating hemorrhage size as total brain volume percentage remains a challenge. Hong and Lee’s study describe ICH in neonates and takes into account the location and type of hemorrhage^8^. This study was a descriptive study, and there is no scale or numerical score to reflect the severity of hemorrhage. The PVHI Score by Bassan et al., has many positive features including location and extent of hemorrhage and a numerical score reflecting severity^11^. However, this scale is limited to PVHI in infants born preterm, and does not fully account for the size of hemorrhage.

Given the limitations of existing pediatric ICH classification, we developed the NeoCVST Score to characterize the location, type and severity of hemorrhage secondary to neonatal CVST. We also wanted to incorporate a numerical score to grade the severity of hemorrhage in neonatal CVST. The NeoCVST Score was applied to our cohort of neonates with CVST and we found that it was able to characterize the full spectrum of ICH and ischemic brain injury. We also found that the NeoCVST total score and subscores for thalamic hemorrhage and ischemia were predictive of neurodevelopmental outcomes. More specifically, we found that increasing NeoCVST total score and subscores for thalamic hemorrhage or ischemia were associated with worse neurodevelopmental outcomes. These results were similar when our analysis was repeated with term neonates, with NeoCVST total score and subscores for thalamic hemorrhage and ischemia being significant predictors of outcome. Interestingly, NeoCVST subscores for GMH-IVH, and hemorrhage in the parenchyma, cerebellum and brainstem, and extra-axial hemorrhage were not associated with neurodevelopmental outcome These results highlight the importance of injury location in CVST, and that involvement of specific brain regions (e.g. thalamus) should be considered for clinical management and prognostication.

The NeoCVST Score is simple and can be easily used for clinical or research purposes. The scale can be applied to neonates in the acute care setting to obtain a standardized description of ICH. We anticipate that the NeoCVST Score will be clinically relevant to Neurologists, Neonatologists and Neuroradiologists to characterize brain injury secondary to neonatal CVST. In terms of broader scale impact, the NeoCVST score can be used for risk stratification and define inclusion/exclusion criteria for randomized controlled trials of anticoagulation therapy in neonatal CVST. Grading the severity of hemorrhage secondary to neonatal CVST based on NeoCVST cutoff scores may help guide treatment decisions for anticoagulation. Anticoagulation treatment for neonatal CVST remains mixed, with some centres favoring anticoagulation even in the presence of ICH, while others choose a more conservative approach with supportive care and follow-up imaging^15^. The hesitancy to anticoagulate in neonatal CVST is often related to the risk of new hemorrhage, or worsening of pre-existing hemorrhage. In this study, a large proportion of neonates were treated with anticoagulation (66%), and most had no anticoagulation related complications. One participant had worsening of extra-axial hemorrhage after starting anticoagulation, and therapy was held for 3 days and then restarted three days later with no further complications. Previous studies from our group have found that anticoagulation in neonatal CVST was safe and associated with better radiological outcome (reduced risk of thrombus propagation)^5,12^. Randomized clinical trials (RCTs) are needed to examine the efficacy of anticoagulation therapy in neonatal CVST, and this knowledge gap has been recognized with calls for multi-centre RCTs for neonatal CVST^16^. The NeoCVST Score can help with inclusion/exclusion criteria for RCTs and discriminate between neonates who have mild hemorrhage from those with more severe hemorrhage.

Strengths of this study include an appropriate sample size of neonates with CVST to capture the full spectrum of ICH seen in this population. Neonates were followed as outpatients allowing for the assessment of long-term neurodevelopmental outcomes. Limitations of the study should be acknowledged. First, the cohort of neonates with CVST included in the study were diagnosed with CVST between 2000-2025. There may have been differences in the approach to diagnosis and management of neonates over these 25 years that we were unable to control for. Second, the NeoCVST Score could not discriminate between causes of intracranial hemorrhage, and there may have been differences in the type and severity of hemorrhage due to comorbidities (e.g. meningitis, coagulopathy). Third, while our models controlled for sex, prematurity and associated brain injury, we could not control for the severity of associated brain lesions, genetic and epigenetic differences and home environment which are all known to influence neurodevelopmental outcomes^5,17–19^. Finally, the NeoCVST score demonstrated moderate-good inter-rater reliability demonstrating measurement reproducibility, and predictive validity, and future studies will need to validate the scale in an external cohort.

In summary the NeoCVST score is a new classification and scoring system to better characterize the spectrum of ICH associated with neonatal CVST. NeoCVST total score and subscores for thalamic hemorrhage and ischemia were predictors of neurodevelopmental outcome. We anticipate that this tool will help guide risk stratification, neuroprognostication and treatment decision-making for neonates with CVST. The NeoCVST is simple to use, and can be easily applied for clinical purposes and help guide management in the neonatal intensive care unit. The NeoCVST Score can also be utilized in RCTs to examine the role of anticoagulation therapy in treating neonatal CVST.

## Data Availability

All data will be made available on request.

## Acknowledgements

We would like to thank Alexandra Linds, Scherazad Musaphir and Sujatha Parthasarathy from the Children’s Stroke Program at the Hospital for Sick Children (Toronto, Canada), for their assistance with data collection for this study.

## Tables

### Instructions

- Rate intracranial hemorrhage in the following locations using the first brain MRI/MRV when CVST was detected, using T1-weighted, T2-weighted or susceptibility weighted (SWI/GRE/MPGR) sequences.
- For hemorrhage: if only thalamic hemorrhage rate in “thalamic hemorrhage” category, if thalamic hemorrhage and multifocal parenchymal hemorrhage rate in “thalamic hemorrhage” **AND** also rate in “parenchymal hemorrhage”.
- For ischemic lesions (on DWI and ADC): if ischemic lesions only in medullary veins or thalamus, rate in corresponding category, if ischemic lesions in medullary veins and white matter, then rate in corresponding category A**ND** also rate in “white matter category”.

## Supplemental Tables

**Supplemental Table 1.** Clinical characteristics of term neonates with CVST (N=77)

| Variable | Frequency (%) or Mean (SD) |
| --- | --- |
| Sex | Males: 62 (81%)<br>Females: 15 (19%) |
| Mean gestational age in weeks (SD) | 38.8 (2.3) |
| Perinatal risk factors | Vacuum or forceps assisted delivery: 18 (23%)<br>Surgery: 15 (20%)<br>Congenital heart disease: 10 (13%)<br>Infection: 10 (13%)<br>Dehydration: 10 (13%)<br>Hypoxic ischemic encephalopathy: 9 (12%)<br>Coagulopathy: 5 (7%) |
| Mean age at CVST diagnosis in days (SD) | 8.5 (6.5) |
| Clinical features at diagnosis | Seizures: 56 (73%)<br>Abnormal neurological exam: 26 (34%)<br>Asymptomatic: 11 (14%) |
| Location of thrombus | Multiple sinuses involved: 46 (60%)<br>Superior sagittal sinus: 15 (19%)<br>Transverse sinus: 12 (16%) |
| Intracranial hemorrhage<sup>b</sup> | Supratentorial hemorrhage: 42 (54%)<br>GMH-IVH grade 1-2: 14 (18%)<br>GMH-IVH grade 3: 11 (14%)<br>Parenchymal: 29 (38%)<br>Thalamic: 14 (18%)<br>Infratentorial hemorrhage: 16 (21%)<br>Cerebellum: 15 (19%)<br>Brainstem: 3 (4%)<br>Extra-axial hemorrhage 27 (35%)<br>Epidural: 2 (3%)<br>Subdural: 20 (26%)<br>Subarachnoid: 6 (8%) |
| Parenchymal brain injury<sup>c</sup> | Ischemic lesion: 54 (70%)<br>Medullary vein distribution: 11 (14%)<br>White matter: 44 (57%)<br>Cystic white matter lesion: 1 (1%)<br>Basal ganglia: 15 (20%)<br>Thalamus: 21 (27%) |
| Mean NeoCVST Score | 9.6 (7.8) |
| Anticoagulation received | No: 25 (32%)<br>Yes: 46 (60%) at diagnosis, 6 (8%) after propagation |
| Mean duration of anticoagulation in months (SD) | 2.7 (1.2) |
| Reason for not anticoagulating (N=25) | Intracranial hemorrhage: 22 (89%)<br>Non-occlusive thrombus: 3 (11%) |
| Worsening hemorrhage with anticoagulation (N=52) | 1 (2%) |
| Recanalization outcome | Full recanalization: 38 (49%)<br>Partial recanalization: 32 (42%)<br>No change: 1 (1%) |
| Mean age at follow-up in years (SD) | 5.9 (4.9) |
| Mean PSOM Score at last follow-up (SD) | 1.1 (1.7) |
| Clinical outcome based on PSOM (N=75) | No impairment: 31 (41%)<br>Impairment: 44 (59%) |

<sup>a</sup>Cause of death: multiorgan failure 1 (1%), leukemia 1 (1%)
<sup>b</sup>Intracranial hemorrhage: some neonates had more than one type of intracranial hemorrhage injury
<sup>c</sup>Parenchymal brain injury: some neonates had more than one type of parenchymal injury

**Supplemental Table 2.** Logistic Regression Models NeoCVST Score and PSOM Clinical Outcomes in term neonates with CVST.

| Univariate Analysis | Odds ratio | 95% CI | P-Value |
| --- | --- | --- | --- |
| NeoCVST total score | 1.1 | [1.0, 1.2] | 0.03* |
| NeoCVST subscore |  |  |  |
| – Supratentorial hemorrhage GMH-IVH grade 1-2 | 1.2 | [0.4, 4.1] | 0.8 |
| – Supratentorial hemorrhage GMH-IVH grade 1-2, bilateral | 1.2 | [0.6, 2.4] | 0.5 |
| – Supratentorial hemorrhage GMH-IVH grade 3 | 2.9 | [0.6, 14.0] | 0.2 |
| – Supratentorial hemorrhage GMH-IVH grade 3, bilateral | 1.8 | [0.8, 4.0] | 0.2 |
| – Supratentorial parenchymal hemorrhage | 1.5 | [1.0, 2.3] | 0.07 |
| – Supratentorial parenchymal hemorrhage, bilateral | 1.3 | [0.7, 2.4] | 0.4 |
| – Supratentorial thalamic hemorrhage | 2.1 | [1.0, 4.2] | 0.04* |
| – Supratentorial thalamic hemorrhage, bilateral | 4.6 | [1.1, 19.3] | 0.04* |
| – Infratentorial cerebellar hemisphere hemorrhage | 0.9 | [0.3, 2.5] | 0.8 |
| – Infratentorial cerebellar hemisphere hemorrhage, bilateral | 1.0 | [0.4, 2.0] | 0.9 |
| – Infratentorial cerebellar vermis hemorrhage | 1.2 | [0.4, 4.1] | 0.8 |
| – Infratentorial brainstem hemorrhage | 1.3 | [0.6, 2.8] | 0.5 |
| – Extra-axial hemorrhage, supratentorial | 0.6 | [0.3, 1.3] | 0.2 |
| – Extra-axial hemorrhage, supratentorial, bilateral | 0.7 | [0.4, 1.5] | 0.4 |
| – Extra-axial hemorrhage, infratentorial | 0.6 | [0.3, 1.5] | 0.3 |
| – Extra-axial hemorrhage, infratentorial, bilateral | 0.7 | [0.4, 1.4] | 0.4 |
| – Ischemic medullary vein distribution | 1.1 | [0.5, 2.1] | 0.8 |
| – Ischemic medullary vein distribution, bilateral | 1.0 | [0.5, 2.1] | 0.9 |
| – Ischemic white matter | 1.2 | [0.8, 2.0] | 0.4 |
| – Ischemic white matter, bilateral | 1.4 | [0.8, 2.3] | 0.2 |
| – Ischemic basal ganglia | 1.3 | [0.8, 2.2] | 0.3 |
| – Ischemic basal ganglia, bilateral | 1.4 | [0.7, 3.0] | 0.4 |
| – Ischemic thalamus | 3.0 | [1.4, 6.5] | 0.004* |
| – Ischemic thalamus, bilateral | 4.7 | [1.6, 14.1] | 0.006* |
| **Multivariate Analysis** | **Odds Ratio** | **95% CI** | **P-Value** |
| Covariates: sex, multi-focal hemorrhage, hypoxic-ischemic encephalopathy |  |  |  |
| NeoCVST total score | 1.1 | [1.0, 1.2] | 0.03* |
| NeoCVST Subscore |  |  |  |
| – Supratentorial thalamic | 2.1 | [1.0, 4.4] | 0.04* |
| – Supratentorial thalamic, bilateral | 4.7 | [1.1, 20.3] | 0.04* |
| – Ischemic thalamic | 3.2 | [1.5, 6.9] | 0.003* |
| – Ischemic thalamic, bilateral | 5.2 | [1.7, 16.4] | 0.005* |

## Notes

### Competing Interest Statement

The authors have declared no competing interest.

### Author Declarations

Institutional Research Ethics Board (The Hospital for Sick Children)

